# The effect of population mobility restrictive measures on the incidence of SARS-CoV-2 infection in the early phase of the pandemic

**DOI:** 10.1101/2021.04.13.21255382

**Authors:** Luis Gustavo Modelli de Andrade, Gustavo Fernandes Ferreira, Helio Tedesco-Silva

## Abstract

**Background:** This analysis aims to assess the association between population restrictive measures and the cumulative number of confirmed COVID-19 cases in the early phase of pandemic.

**Methods:** We compared mobility data extracted from the Mobility Reports provided by Google with the cumulative number of confirmed cases of COVID-19 of 15 countries provided by John Hopkins University. We compered the number of confirmed COVID-19 cases before and after the peak effect (PE) of population mobility restrictions in each country, defined as the highest percent reduction in mobility measurements.

**Results:** Time to PE of population mobility restrictions ranged between 16 and 45 days after the report of the index COVID-19 confirmed case in each country. The most frequent reductions in activities were retail & recreation, parks, and transit & stations, ranging from 30% to 90%. Despite this variability in PE among the countries, the predicted smooth effect after the PE of population mobility restrictions was observed in almost all countries.

**Conclusions:** These data suggest that the reduction in mobility was associated with a decrease in the cumulative total number of COVI-19 cases in each country, underscoring that the use of widely available real-time surveillance data might be a valuable resource during this pandemic.

## Introduction

The coronavirus disease 2019 (COVID-19) pandemic represents a global public health emergency. As of April 2020, a total of 1.690.000 confirmed cases and over 100.000 deaths have been reported [1]. The lack of effective pharmacological treatment or vaccines underscores the central role of population mobility restrictions, as early implemented measures such as social distancing were fundamental to reduce viral spread [2]. Public data is another tool that may assist in decision making and in monitoring the effectiveness of community measures. Published data used anonymized public mobile phone data to measure the spread of malaria [3] and Ebola [4].

The use of Google trends data may be potentially helpful in defining the timing and location for implementing appropriate changes in population mobility. Also, this tool can evaluate the association between population mobility restrictions and the number of confirmed COVID-19 cases [5].

We then sought to test the hypothesis that a reduction in community mobility was associated with a slower progression of the COVI-19 epidemic. Therefore, this analysis assessed the effectiveness of government implemented restrictions in community mobility on the observed number of confirmed COVID-19 cases in the early phase of pandemic.

## Material and Methods

### Confirmed COVID-19 Cases Data

We obtained the number of confirmed cases from the COVID-19 Dashboard operated by the Center for Systems Science and Engineering (CSSE) Johns Hopkins University (JHU) [6]. The data set is publicly available and provides the number of confirmed cases by day and country.

### Community Mobility Reports Data

The changes in population mobility in each country were obtained from the Community Mobility Reports [7]. The reports use aggregated, anonymized data to chart movement trends over time by geography, across different high-level categories of places such as retail and recreation, groceries and pharmacies, parks, transit & stations, workplaces, and residential. These categories captured access to essential services and were useful to measure the social distancing efforts. We grouped into six categories; recorded the variables in percentage point increase or decrease in visits:

1. Retail & recreation: Mobility trends for places like restaurants, cafes, shopping centers, theme parks, museums, libraries, and movie theaters.
2. Grocery & pharmacy: Mobility trends for places like grocery markets, food warehouses, farmers’ markets, specialty food shops, drug stores, and pharmacies.
3. Parks: Mobility trends for places like national parks, public beaches, marinas, dog parks, plazas, and public gardens.
4. Transit stations: Mobility trends for public transport hubs such as subway, bus, and train stations.
5. Workplaces: Mobility trends for places of work.
6. Residential: Mobility trends for places of residence.

### Maximum Reduction in Mobility (lowering population mobility)

We defined the peak effect (PE) of population mobility restrictions as the highest percent reduction in community mobility measurements in one of the five mobility categories (Retail & recreation, Grocery & pharmacy, Parks, Transit stations, or Workplaces).

### Study Period and Countries

The study period started on 2020-02-15 when the data community mobility reports became available and ended on 2020-04-11 [7]. We included in these analysis countries that had mobility data available [7] and had a smartphone penetration of more than 50% [8]. The smartphone penetration is the ratio of smartphone users by the total population. We defined for each country, day zero, as the first confirmed COVID-19 case reported.

### Statistical Analysis

The cumulative log incidence of daily total confirmed COVID-19 cases by 100,000 persons (100K) and the daily percent changes in the six categories of the community mobility reports were plotted from the index case in each country. After an exploratory inspection, plots of data we group the 15 countries in clusters. Countries with a similar trend in community mobility reports were grouped for comparison, based on visual inspection of the plots, were analyzed in clusters.

The association of population mobility restrictions and the incidence of COVID-19 cases observed in each country were determined using two different models. First, we fitted an autoregressive generalized additive model (gam) using the cumulative log total confirmed COVID-19 cases as the outcome and day as a predictor (smoothing parameter) [9-10]. Because daily log total confirmed COVID-19 cases are highly correlated with previous values in this time series, we used an autoregressive model (gamm) that combines a gam model with a mixed effect model. We compared the smoothing effect observed 15 before and 15 days after the PE separately. Then we plotted daily projected log total COVID-19 cases 15 days before and 15 days after peak effect of population mobility restrictions and compared them with the observed cases (black) in each country. The 15-days timeframe was defined by the median time to reach PE and was a reasonable short period to evaluate changes in the incidence of COVID-19. To measure the smooth effect, we assess the values of estimated degrees of freedom (EDF). Values of EDF close to one suggest that the model approximates a linear effect, and higher EDF values indicate more complex splines. More significant amounts of EDF after PE imply a “smoothing effect,” confirming a reduction in the incidence of the number of confirmed cases (fattening the curve of total cases). Besides, we compared using ANOVA the difference in EDF before and after PE.

To further confirm that any of the mobility changes were associated with reduced cumulative log total COVID-19 cases, we also fit a global model using a Bayesian approach. In this model, we included all the 15 countries as a random effect, daily mobility changes as a factor, log cases as the outcome, and the day as the smooth factor. We used a gam model fitted with Stan estimated using Markov Chain Monte Carlo (MCMC) sampling with four chains of 2000 interactions and a warm-up of 1000. We considered weakly informative priors to the joint probability of posterior distribution. We found the median as point estimative and 90% highest density interval (HDI) to evaluate the effect of predictors. Finally, to explore potential leads and lags associated with the observed PE time, we also fitted gamm models using different time points (5, 10, 15, and 20 days) before and after the peak effect of population mobility restrictions. We analyzed with R version 3.6.3 [11] with the packages ggplot2, mgcv, and stanarm.

## Results

We identified 15 countries with mobility data available and more than 50% mobile phone penetrance (Argentina, Australia, Brazil, Canada, Germany, Spain, France, United Kingdom, Italy, Japan, South Korea, Mexico, Sweden, Turkey, and the United States.) China, Iran, and Russia did not have mobility data available and African, and some Asian countries (India) did not have mobile phone penetrance higher than 50%.

The maximum percentage reduction in each of the mobility categories for each country is shown in Table 1. Countries are listed based on the total number of confirmed COVID-19 cases per 100,000 persons (100K), beginning with Spain with 347/100K to Mexico with 3/100K. There was a high variability among the magnitude of the PE of population mobility restrictions, both among the categories of community mobility and among the countries. The time to reach the PE varied from 16 days in the United States and 45 days in Japan. Mobility trends were consistently reduced for retail & recreation, grocery & pharmacy, parks, transit stations, and workplaces; they increased in places of residence, confirming the effectiveness of the strategies. Percent reductions in most mobility categories were high in Spain, Italy, and France and relatively low in Japan, South Korea, Sweden, and the United States. There was an association between the number of SAR-CoV-2 tests per 100K and the number of confirmed COVID-19 cases (r=0.77, p=0.001). There was no association between the time to reach the PE and the maximum reduction in community mobility with the total COVID-19 cases per 100K in each country (Figure 1).

**Table 1:** Effect of mitigation strategies on maximum percent reduction in the mobility categories. Time to PE: time elapsed from the first confirmed COVID-19 case to the highest percent reduction in one of the five mobility categories.

| **Country** | **Retail & Recreation (%)** | **Transit & station (%)** | **Grocery & pharmacy (%)** | **Parks (%)** | **Residential (%)** | **Workplace (%)** | **Time to PE (days)** | **Cases Per 100K** | **Tests Per 100k** | **Total Cases** |
| --- | --- | --- | --- | --- | --- | --- | --- | --- | --- | --- |
| **Spain** | -96 | -91 | -90 | -89 | +42 | -88 | 28 | 347 | 6471 | 163027 |
| **Italy** | -95 | -86 | -87 | -90 | +35 | -71 | 28 | 252 | 5224 | 152271 |
| **France** | -90 | -87 | -73 | -80 | +22 | -72 | 30 | 142 | 2066 | 94863 |
| **Germany** | -81 | -70 | -92 | -44 | +28 | -85 | 42 | 150 | 4331 | 124908 |
| **United States** | -43 | -39 | -15 | -16 | +36 | -47 | 16 | 160 | 4328 | 526233 |
| **United Kingdom** | -84 | -70 | -45 | -53 | +31 | -78 | 39 | 120 | 4637 | 79874 |
| **Sweden** | -47 | -52 | -22 | -38 | +20 | -79 | 40 | 99 | 2051 | 10151 |
| **Canada** | -72 | -75 | -52 | -36 | +34 | -78 | 42 | 62 | 3657 | 23303 |
| **Turkey** | -92 | -90 | -83 | -81 | +32 | -78 | 22 | 64 | 2068 | 52167 |
| **Australia** | -76 | -75 | -54 | -51 | +33 | -80 | 42 | 25 | 4550 | 6303 |
| **South Korea** | -41 | -36 | -18 | -38 | +13 | -16 | 20 | 20 | 1527 | 10480 |
| **Brazil** | -75 | -72 | -44 | -68 | +24 | -70 | 25 | 10 | 352 | 20727 |
| **Japan** | -24 | -44 | -4 | -32 | +33 | -52 | 45 | 5 | 205 | 6005 |
| **Argentina** | -89 | -86 | -64 | -87 | +16 | -74 | 26 | 4 | 252 | 1975 |
| **Mexico** | -71 | -69 | -31 | -56 | +29 | -70 | 32 | 3 | 147 | 3844 |

**Figure 1:**
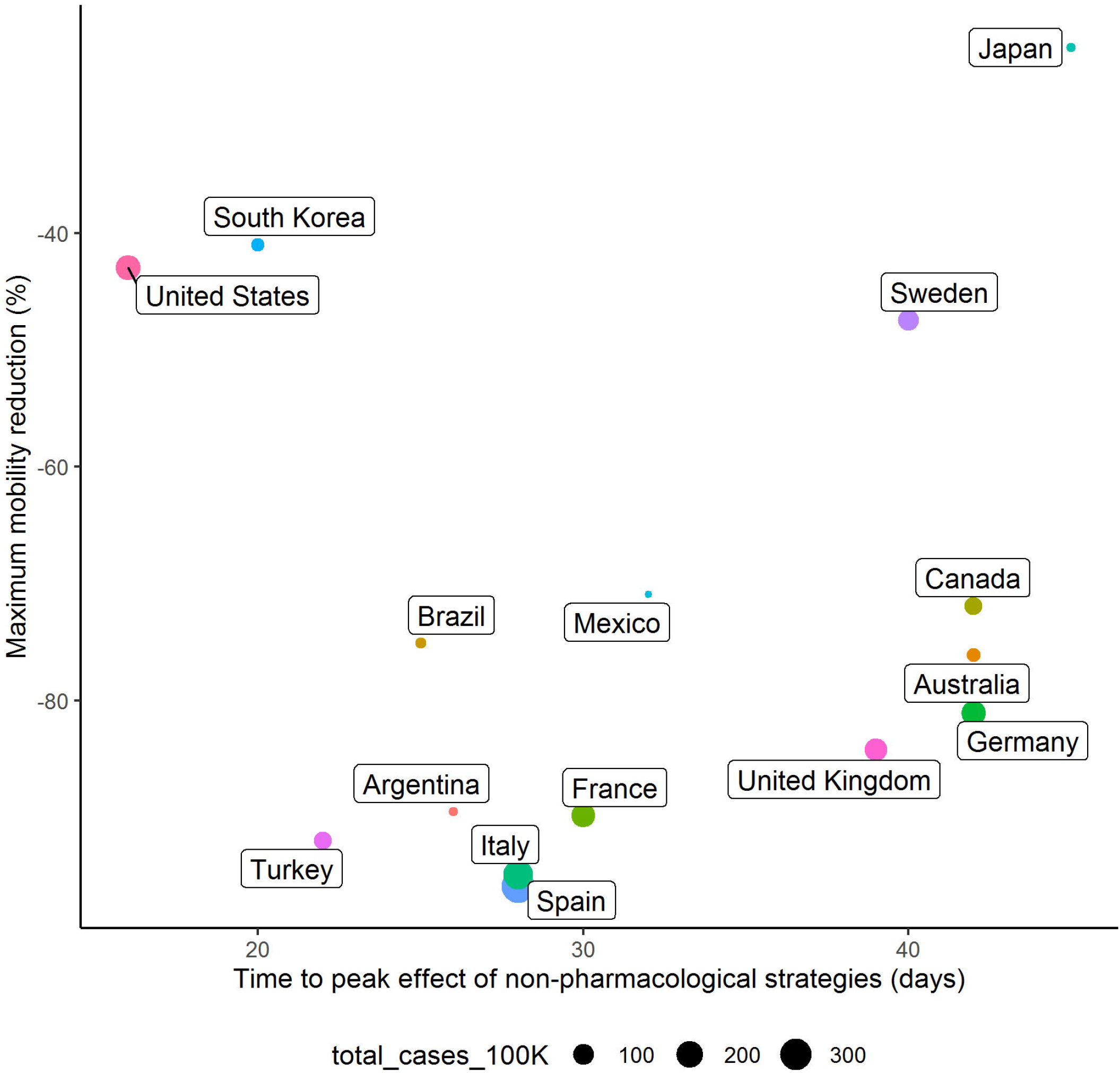
Association between the timing (day) and the maximum reduction in mobility (%) achieved in each country. The size of each dot is proportional to the total number of COVID-19 cases by 100k in each country.

The cumulative incidence of log total confirmed COVID-19 cases and the percent changes in all six community mobility categories since the first confirmed COVID-19 case in each country are shown in Figures 2-8. Visual inspection of the curves suggests that the incidence of the total log COVID-19 cases decreased after reaching the PE of population mobility restrictions, with exception of Japan and Sweden.

**Figure 2:**
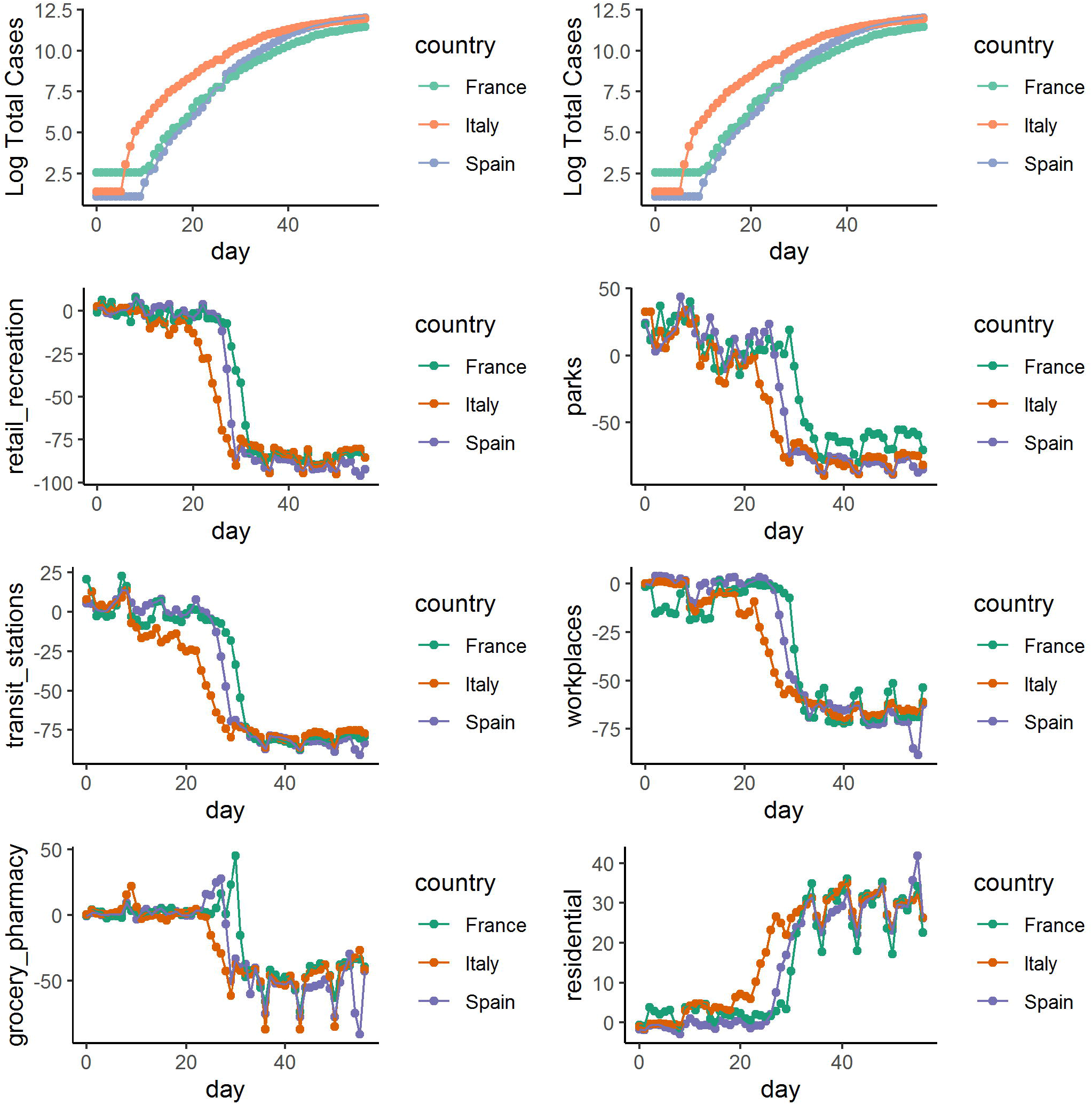
Daily changes in log total COVID-19 cases and mobility reports in group 1 (France, Italy, and Spain)

**Figure 3:**
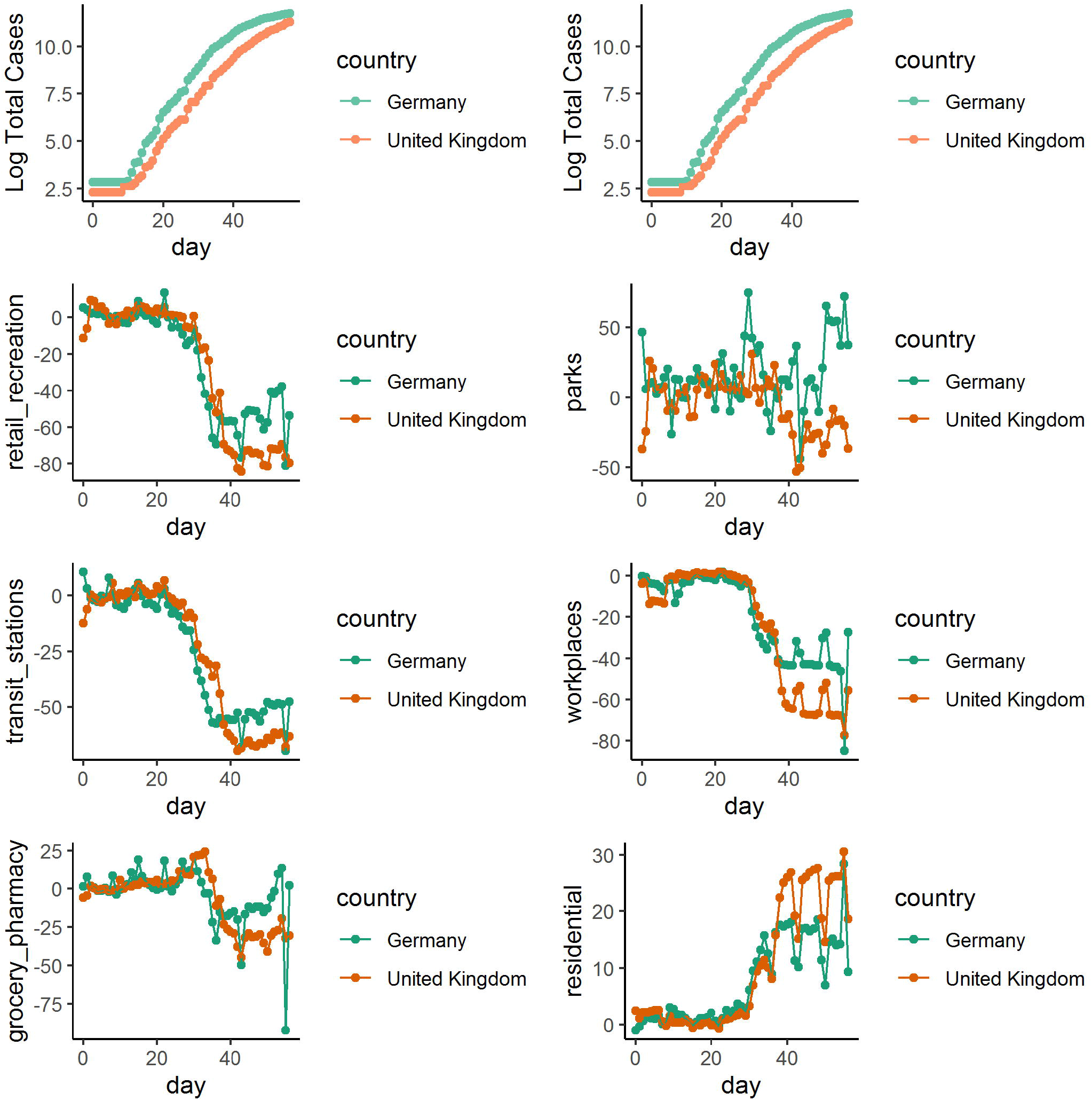
Daily changes in log total COVID-19 cases and mobility reports in group2 (Germany and United Kingdom).

**Figure 4:**
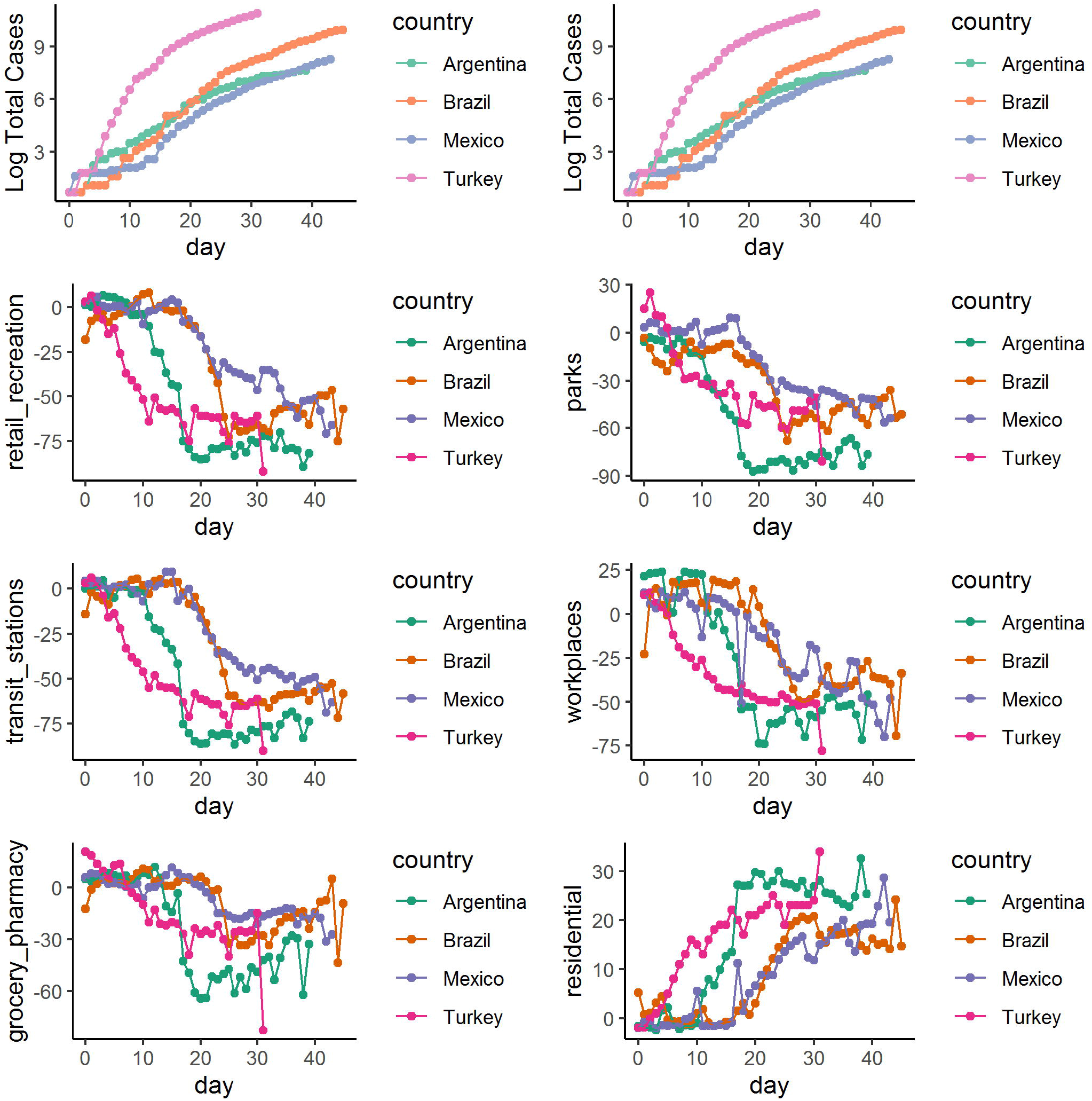
Daily changes in log total COVID-19 cases and mobility reports in group 3 (Argentina, Brazil, Mexico, and Turkey)

**Figure 5:**
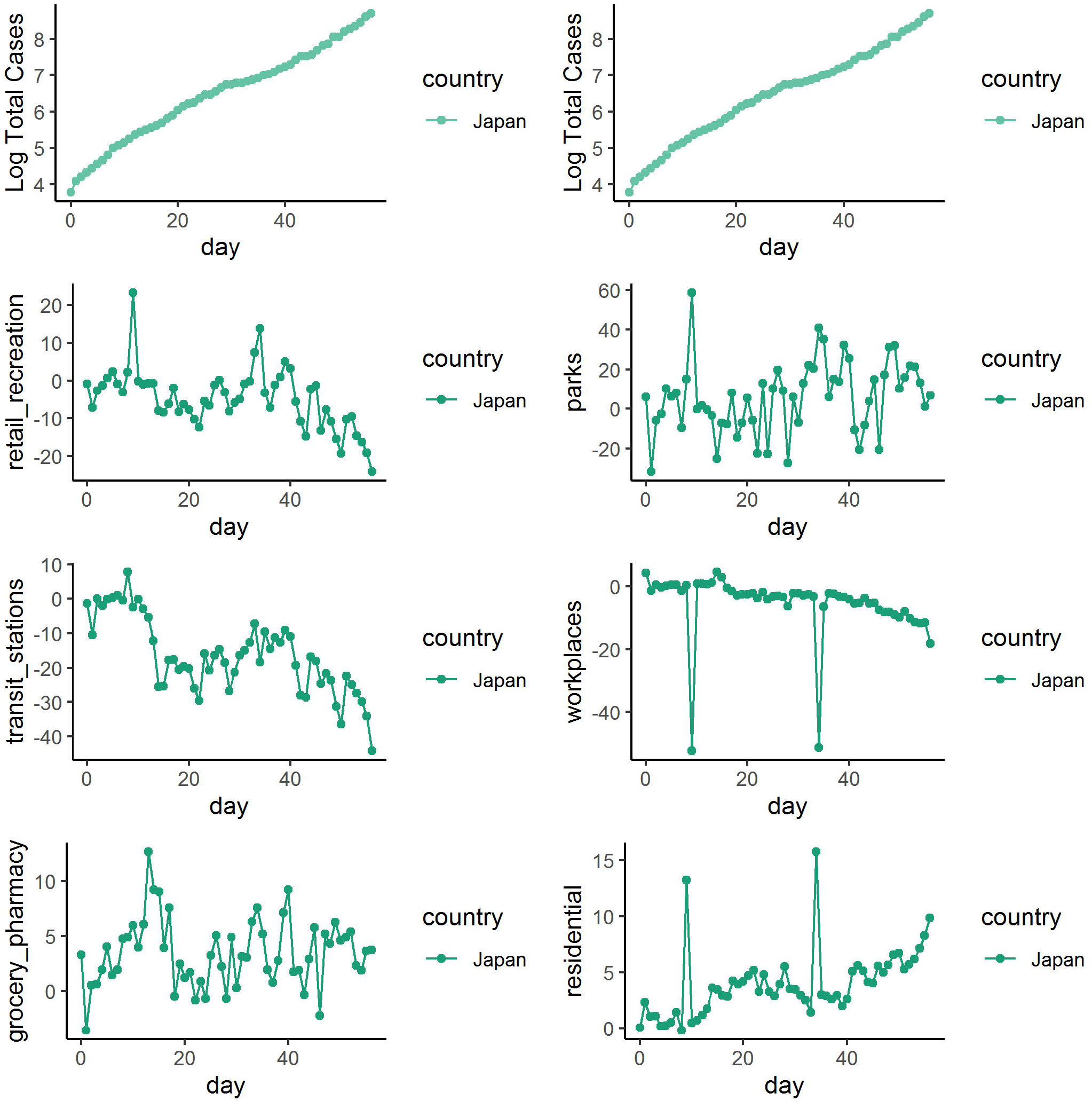
Daily changes in log total COVID-19 cases and mobility reports in group 4 (Japan)

**Figure 6:**
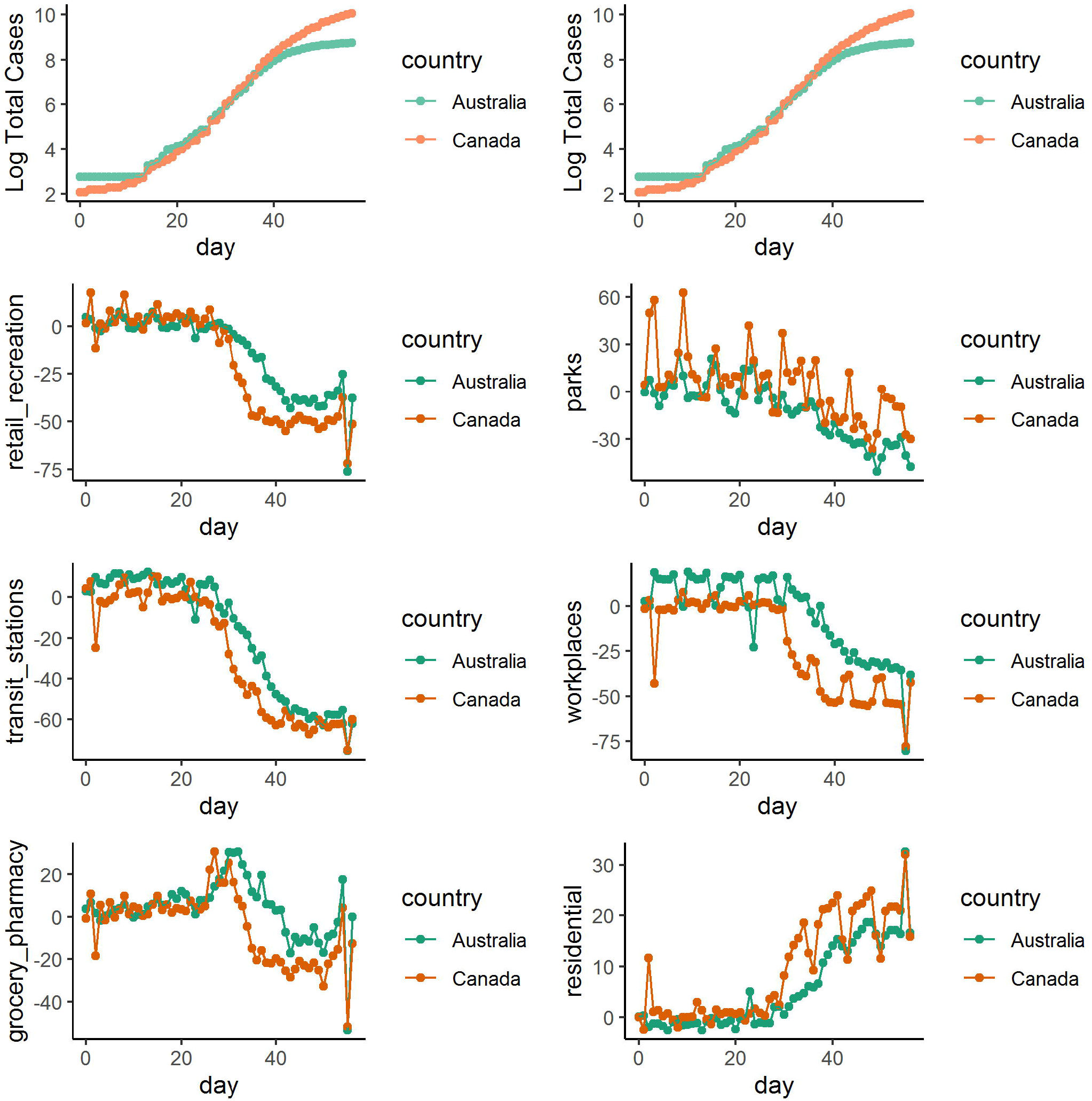
Daily changes in log total COVID-19 cases and mobility reports in group 5 (Australia and Canada)

**Figure 7:**
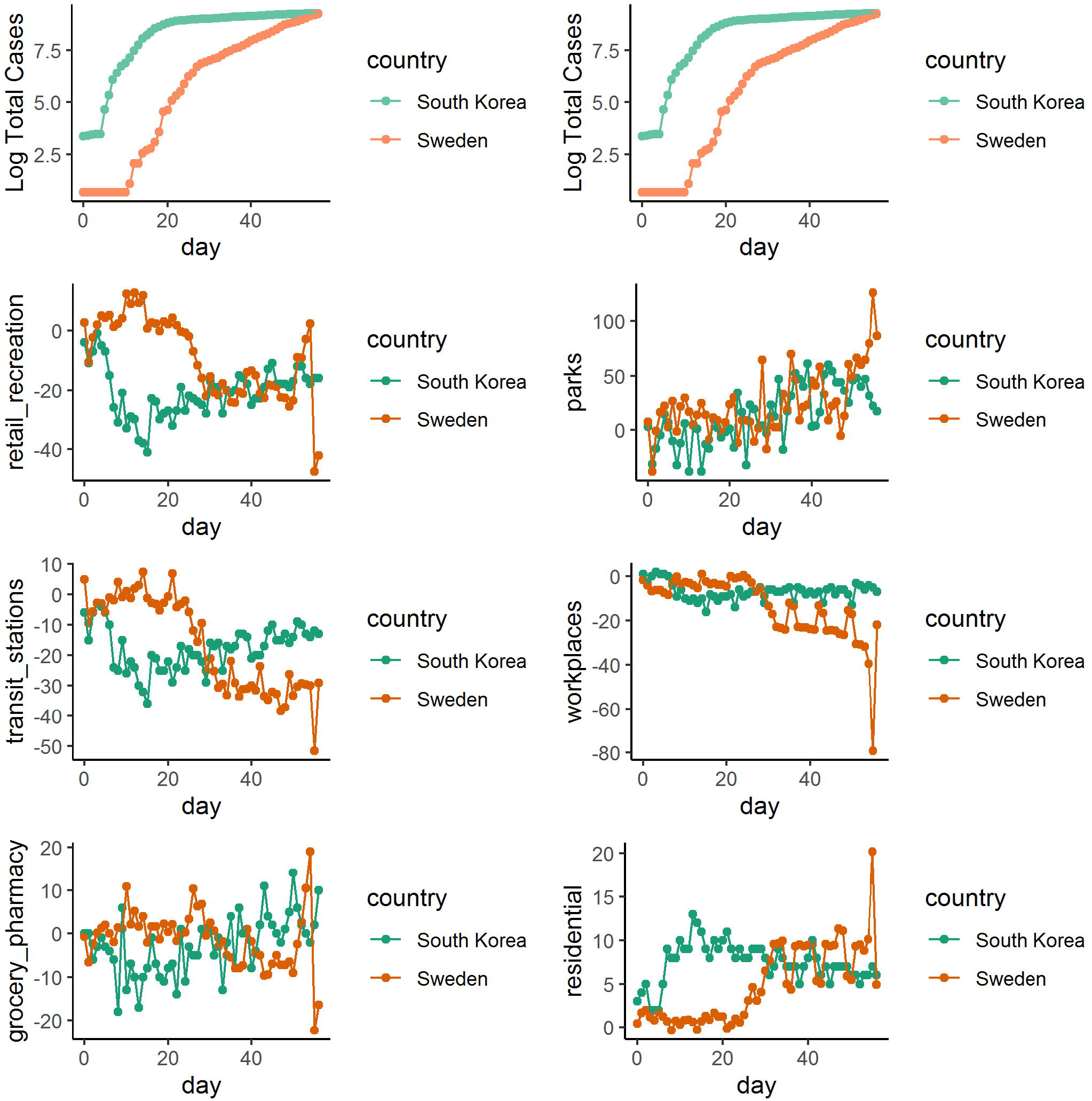
Daily changes in log total COVID-19 cases and mobility reports in group 6 (South Korea and Sweden)

**Figure 8:**
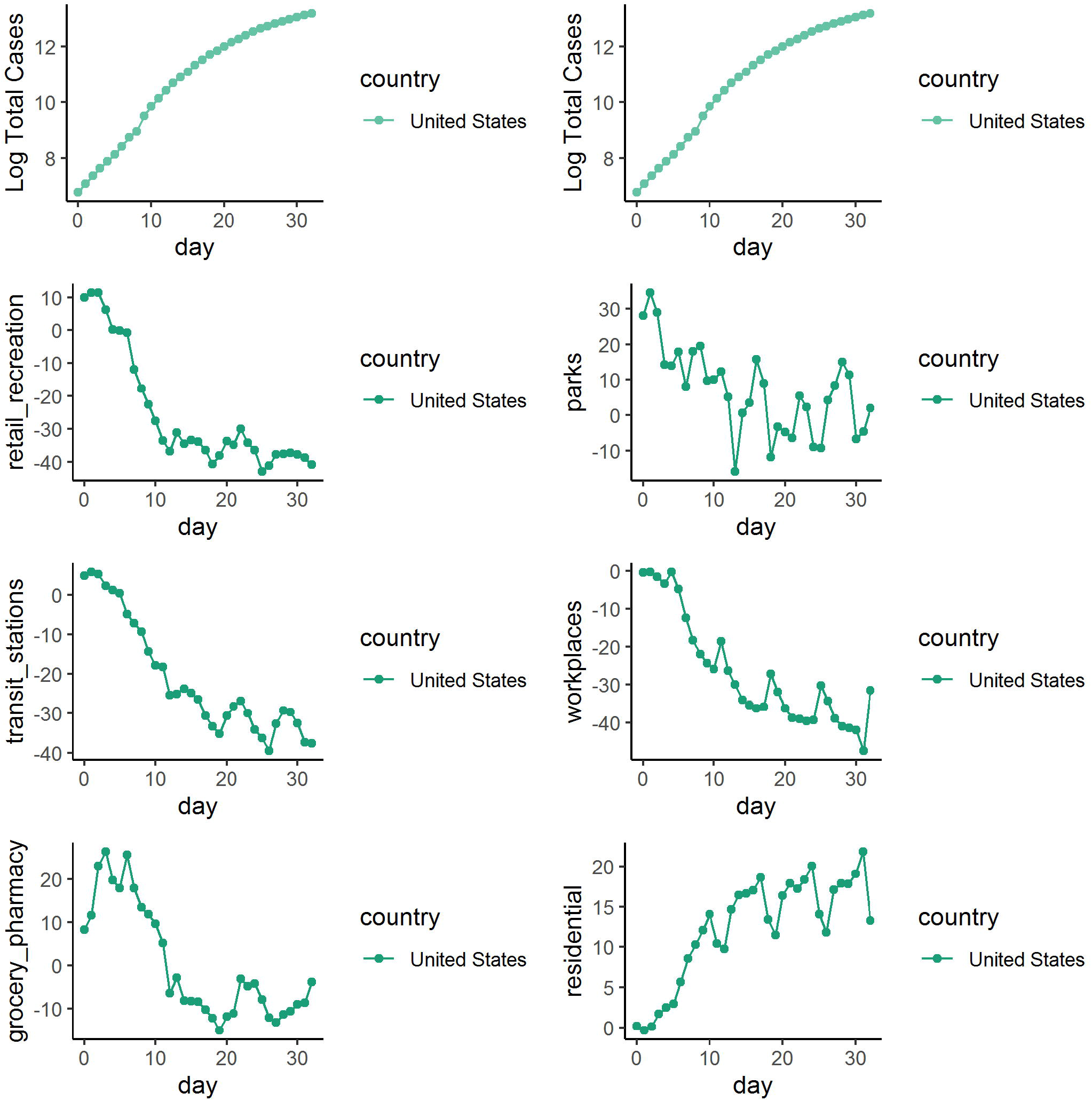
Daily changes in log total COVID-19 cases and mobility reports in group 7 (United States)

To confirm this observation, we compared the smooth effect on log total confirmed COVID-19 cases 15 days before and 15 days after reaching the PE in each country using the gamm model. As an example, the calculated smooth effects 15 days before and 15 days after reaching the PE in Spain is shown in Figure 9.A. Daily log total COVID-19 observed cases followed initially the projections based on the 15 days before, changing afterwards according to the projection based on the data from 15 days after the PE of population mobility restrictions (Figure 9.B.). Similar data for the other countries are shown in Supplementary Figure 1.

**Figure 9.**
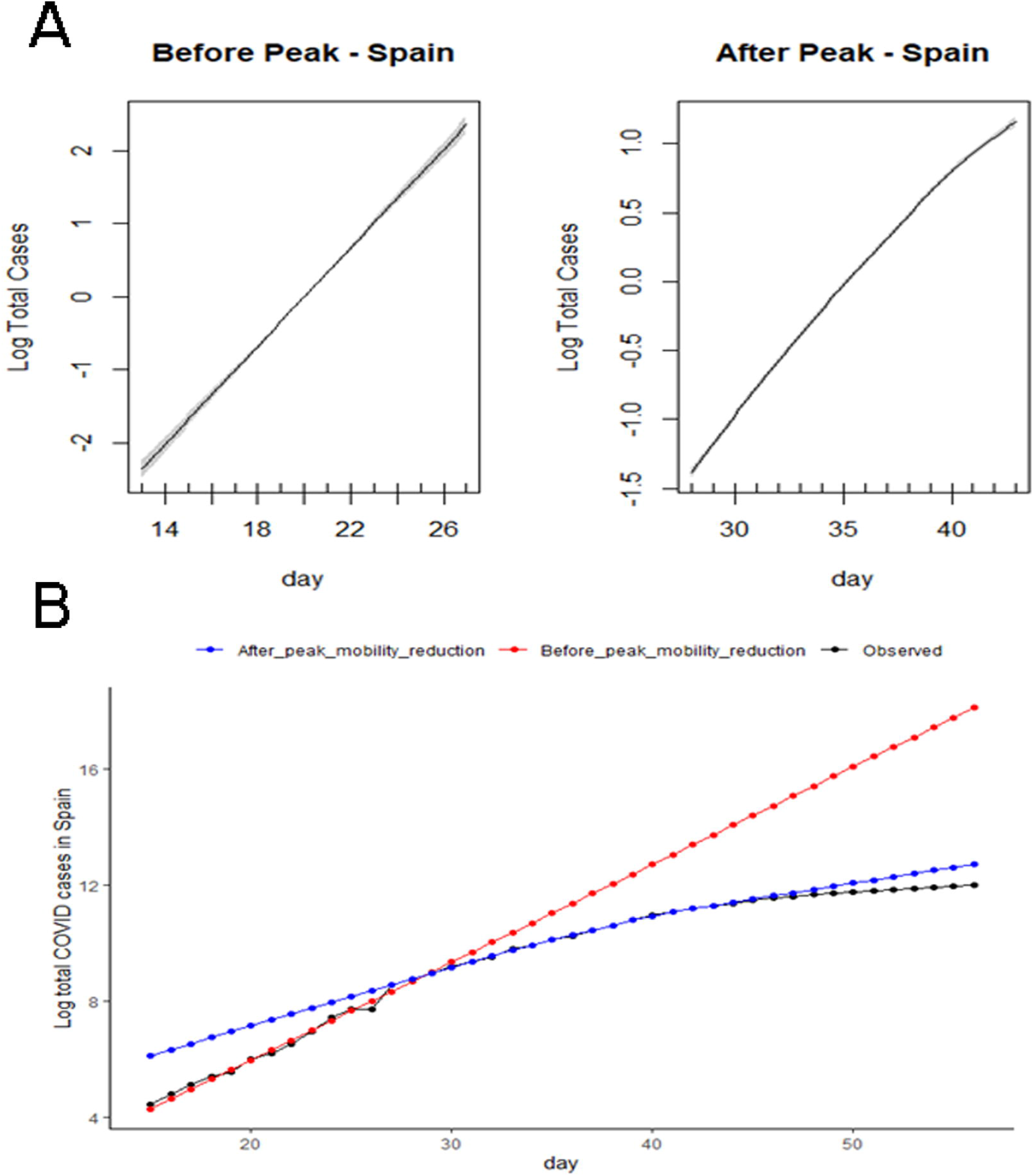
(A) Calculated smooth effects 15 days before and 15 days after reaching the PE in Spain. (B) Daily log total COVID-19 observed cases followed initially the projections based on the 15 days before (red), changing afterwards according to the projection based on the data from 15 days after the PE of lowering population mobility (blue) in Spain.

These analyses were confirmed by the corresponding increase in the values of estimated degrees of freedom (EDF) of the smoothing terms in each country (Table 2). All countries with a consistent reduction of PE (more than 40%) observed a reduction in the incidence of total COVID-19 cases. In Japan and Sweden, the reduction in the log cases was not sustained over time, and consequently, the flattening effect was not observed (Supplementary Figure 1) and no significant difference in EDF was confirmed (Table 2).

**Table 2:**
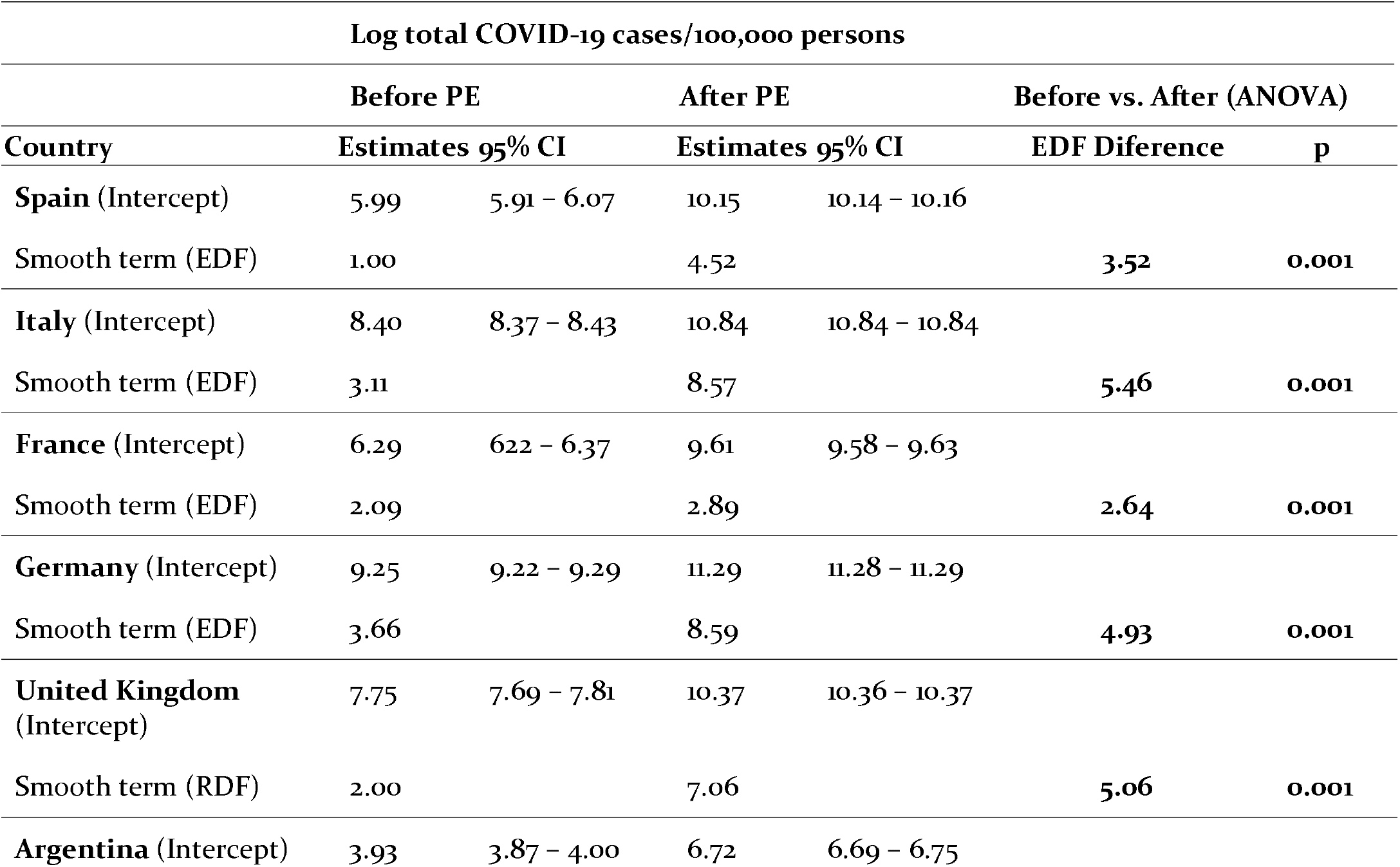
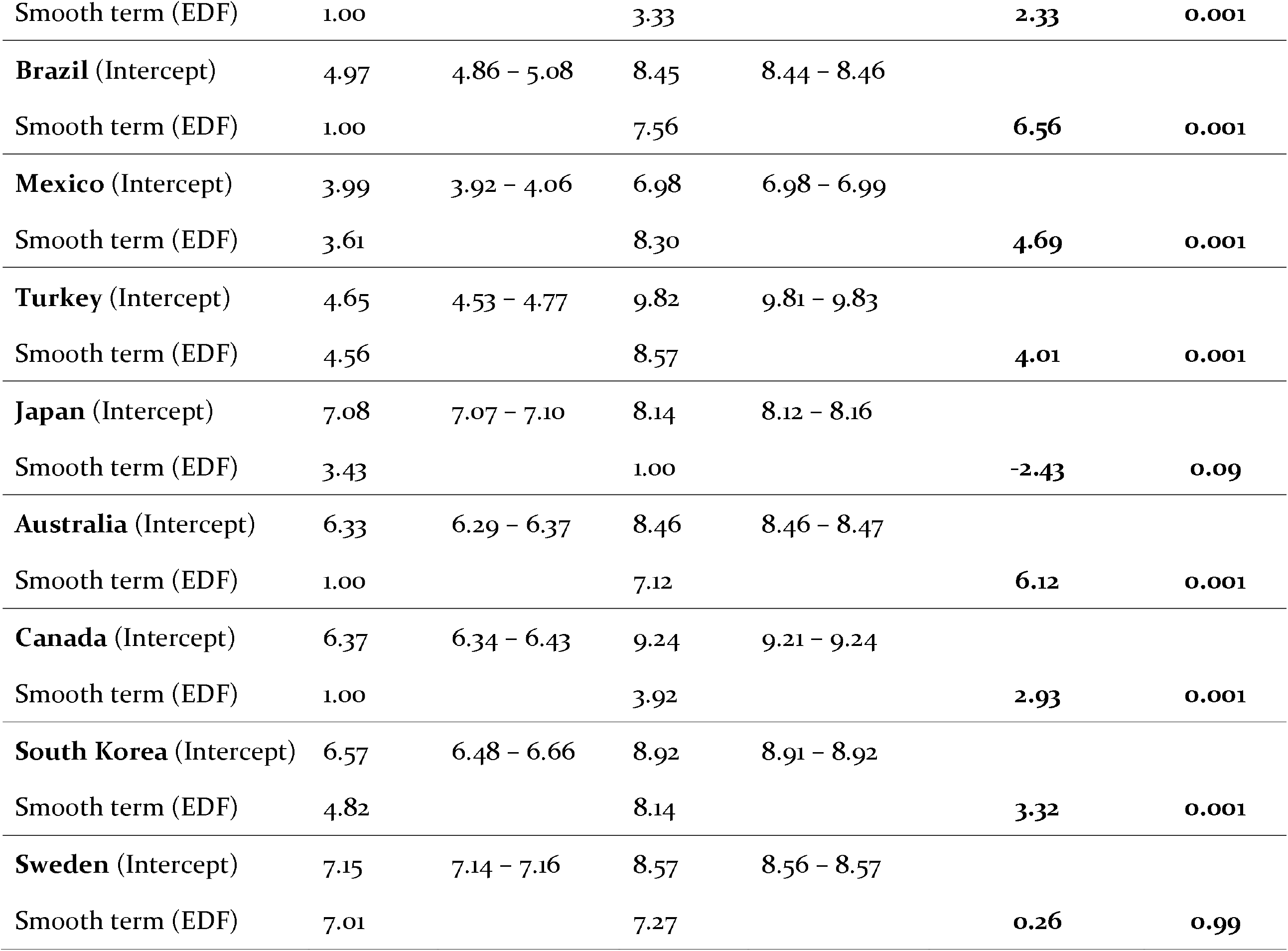
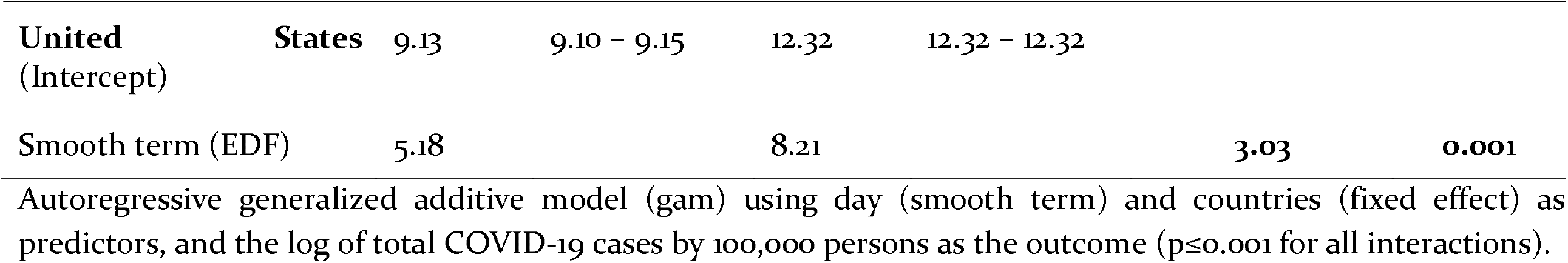
Comparison of the estimated degrees of freedom (EDF) on the prediction of log total COVID-19 cases/100,000 persons, before and after the peak effect (PE) of mitigation strategies on community mobility measurements.

To evaluate if the effect of any percent change in mobility was associated with the cumulative incidence of log cases, we fitted a Bayesian gam model to each of the 15 countries. Every percentage point reduction in mobility resulted in −0.036; 90% CI (HDI_-0.039_, HDI_-0.033_) reduction in log total COVID-19 cases (Table 3).

**Table 3.**
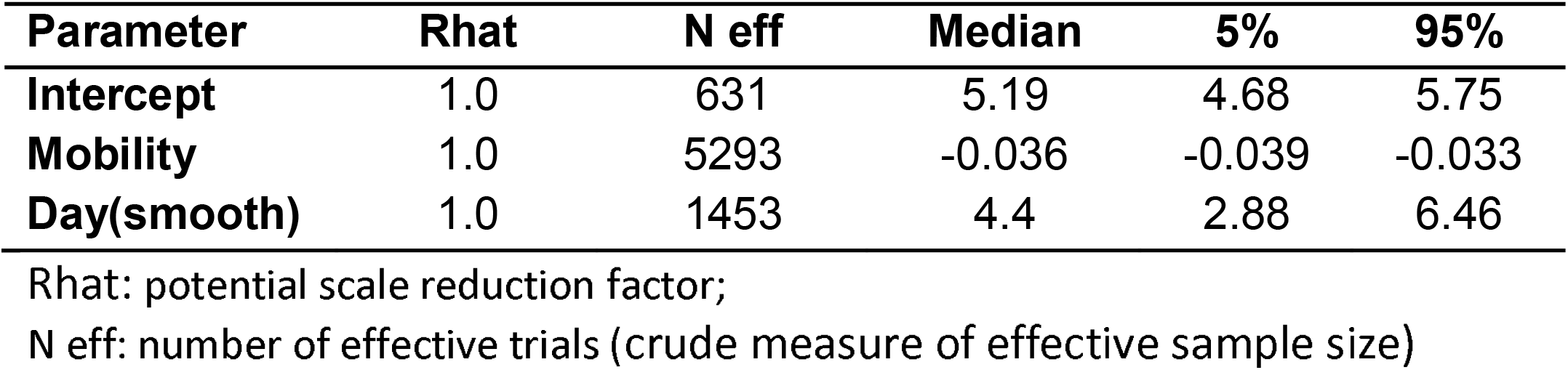
Gam model fitted with Bayesian approach to the log total COVID-19 cases.

Finally, to explore potential leads and lags associated with the observed PE time, we also fitted gamm models using different time points (5, 10, 15, and 20 days) before and after the peak effect of population mobility restrictions. These analyses did not change the interpretation of the data (data not shown).

## Discussion

In this study we evaluated the community mobility reports during the COVID-19 pandemic in 15 different countries. The analysis showed that the implemented population mobility restrictions were effective in reducing the number of COVID-19 cases (flattening the incidence curve), which has been previously supported only by empirical mathematical models [2, 12]. Therefore, real-time community mobility reports [7] are an effective way to evaluate compliance and effectiveness of government derived population mobility restrictions [12, 13]. Perhaps, this is the first analysis that uses Google community mobility reports to obtain data, verify the effectiveness of implemented measures, and to provide insights for the management of a world public health issue.

The peak of the effect of community measures ranged from 16 to 45 days after the first confirmed case in each country. Maximal reductions in community mobility were observed in retail & recreation, parks, and transit & stations, but ranged from 30% up 90% among the countries. There was no association between the timing of implementation after the first confirmed COVID-19 case and the level of adherence to population mobility restrictions with the total number of COVID-19 cases in each country. This association is influenced by several country-related factors, including the timing of the initiation of the local spread of the virus, the population density, and the ability to perform diagnostic tests, factors that we did not include in our analysis [1]. Nevertheless, we demonstrated that reaching the peak effect was associated with a smoothing effect (flattening) on the incidence of total COVID-19 cases. Our data also suggested that any percent of changes in mobility were associated with a reduction in total COVID-19 cases.

Visual inspection of plots allowed us to identify similar patterns among several countries. We speculate that timing of initiation of mobility restrictions and the rate and the maximal mobility reduction achieved were similar among countries within the same cluster. Japan and Sweden showed an unreliable and low reduction in community mobility reports and consequently did not experience significant reduction in the incidence of COVID-19 cases. This observation reinforces that only a sustained and consistent reduction in mobility is associated with the smoothing effect in the number of COVID-19 cases. Yet, despite lower reductions in mobility, Japan and South Korea showed relatively low cumulative incidence of COVID-19 cases, suggesting that other social and cultural factors, not accounted in this analysis, may also contribute to the spread of the virus.

The population mobility restrictions adopted but different countries, such as the border closure, quarantine, and lockdown maybe captured indirectly by tracking people’s mobility using mobile phones [4, 13] as initially proposed during the epidemic of Ebola. These analyses also provide a better quantification of the mitigation interventions [4, 14]. Different from the outbreak of Ebola that used data from individual mobile phones operator, we use data provided by Google. The Google community reports are de-identified *aggregate* flows that can be computed by phone sensors location. These technologies permit analyzing population mobility while preserving privacy and are rapidly and widely available [15]. The use of digital technology during epidemics has been increasing, and maybe one of the critical factors involved in the rapid containment of the spread of the SARS-CoV-2 in China and South Korea. China uses big data together with government measures, community vigilance, and citizen participation [16].

The interpretation of the our data is limited to countries where mobility reports were available. Therefore, China, Iran, and Russia, were excluded as well as other countries with low mobile phone penetrance, such as some African, Asian, and South American countries.

Also, it is not possible to verify the local effectiveness of population mobility restrictions in each country. The differences in test per 100k may influence the incidence of total COVID-19 cases, limiting a direct comparison between countries. Additionally, other risk factors not included in this analysis may explain the association between peak mobility reduction and the number of COVID-19 cases. This study was limited by a timeline endpoint (April 2020) that antecedes the peak of transmission of COVID-19 in several countries especially in Africa and South America. Also, the data antecedes the second wave of transmission of COVID-19 that affected Europa and the USA.

In summary, this analysis showed that consistent reduction in community mobility was associated with a significant reduction in the cumulative number of COVID-19 cases. The use of real-time surveillance is an underused and powerful tool available to assist governments in dealing with complex public health programs.

## Data Availability

The data set used is publicly available

https://github.com/CSSEGISandData/COVID-19

https://www.google.com/covid19/mobility/

## Abbreviations

100K: 100,000
EDF: estimated degrees of freedom
HDI: highest density interval
GAM: generalized additive model
MCMC: Markov Chain Monte Carlo

## Funding

This research did not receive any specific grant from funding agencies in the public, commercial, or not-for-profit sectors

## Contributions

de Andrade LGM: Study design, Data analysis; Ferreira GF; Data analysis; Tedesco-Filho H: Writing and Data analysis.

## Conflict of Interest

None declared

## Ethical Approval

The data set is publicly available, and ethics was not required

## Notes

### Competing Interest Statement

The authors have declared no competing interest.

### Author Declarations

The data set is publicly available, and ethics was not required

