## Supplementary Figure 1 for "The effect of population mobility restrictive measures on the incidence of SARS-CoV-2 infection in the early phase of the pandemic"

**Figure 1:** Log COVID-19 cases per 100k before and after peak effect of measures (gam model) (A) and daily log total COVID-19 predicted cases before (red) and after (blue) peak effect of measures compared to the observed cases (B) in each country.

**Spain**

(A)

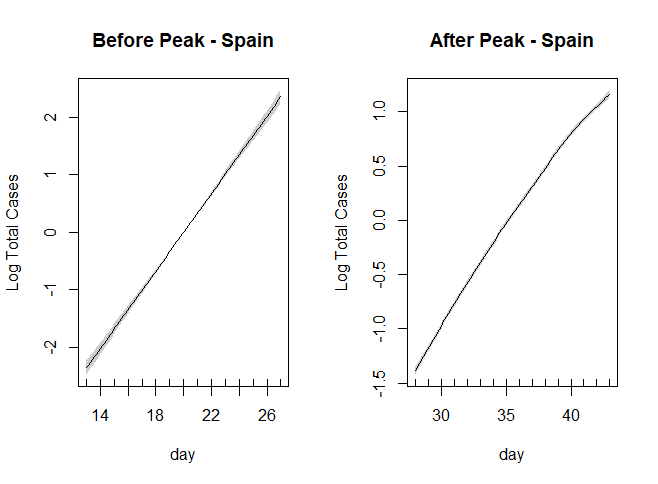

(B)

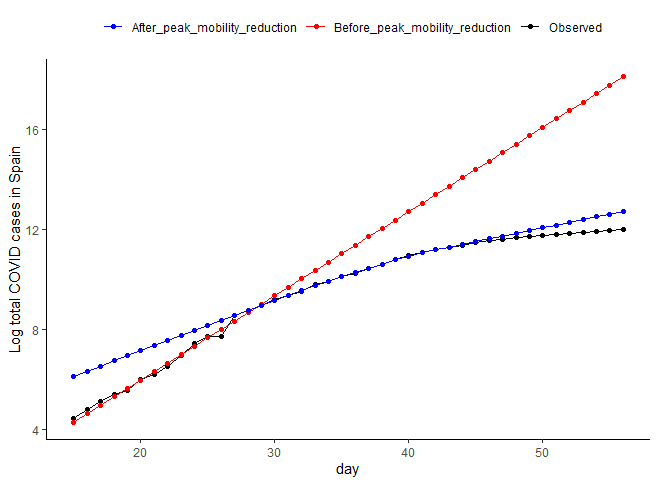

**Italy**

(A)

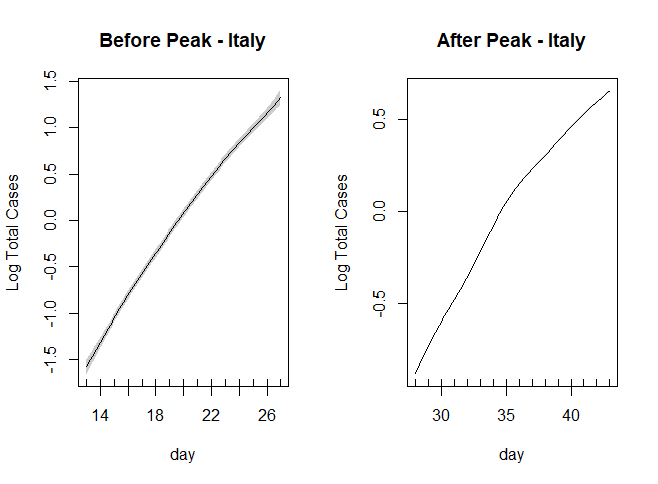

(B)

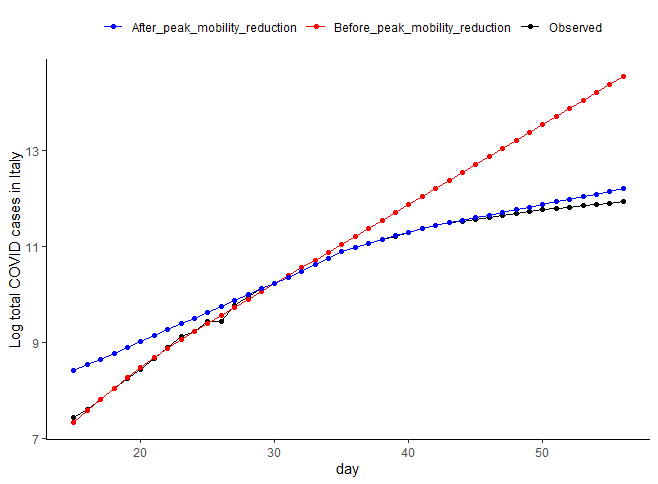

**France**

(A)

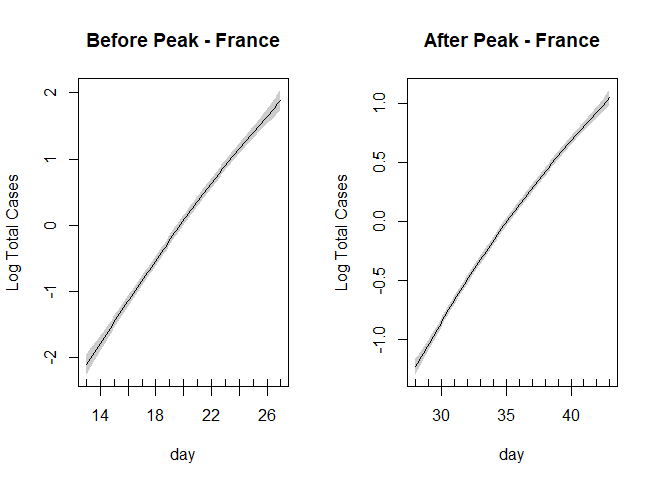
 (B)
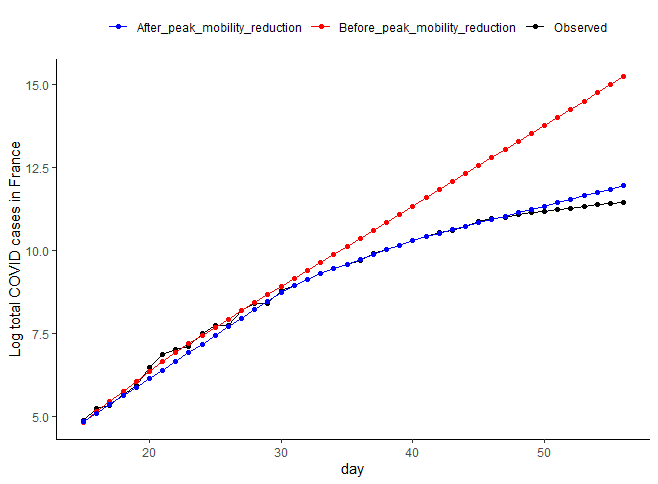

**Germany**

(A)
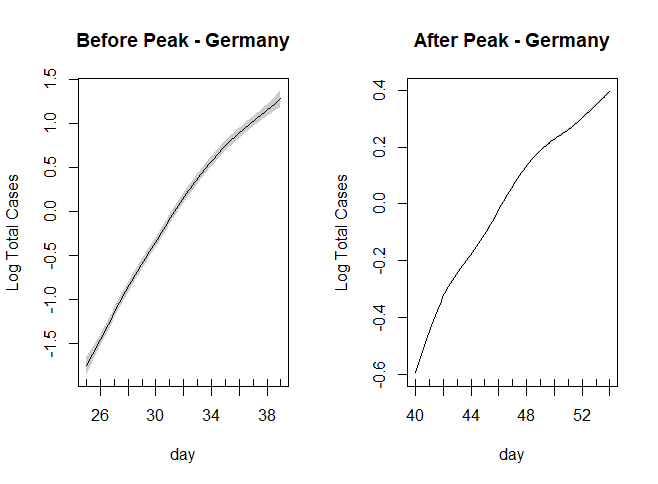
(B)

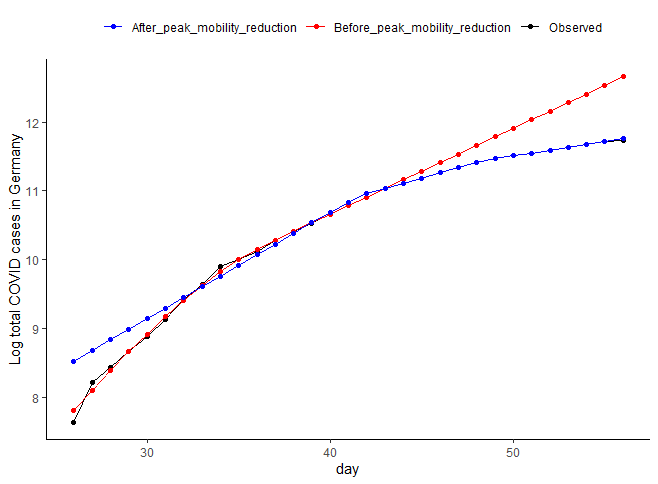

**United Kingdom**

(A)
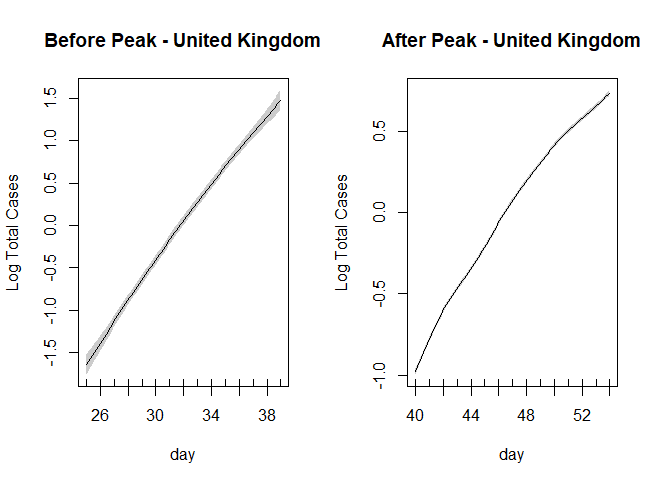
(B)

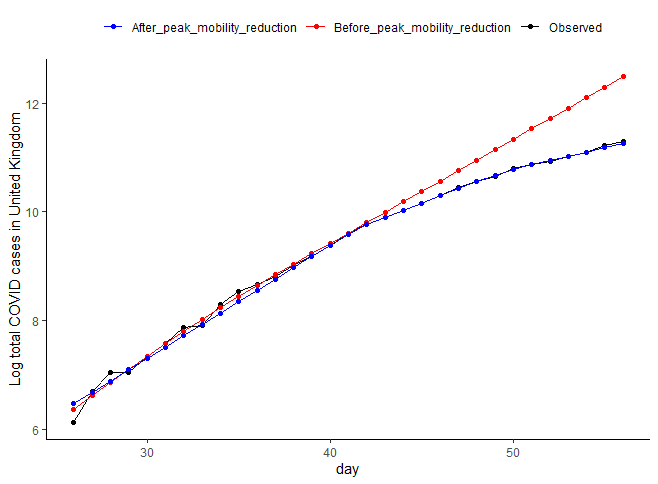

**Argentina**

(A)

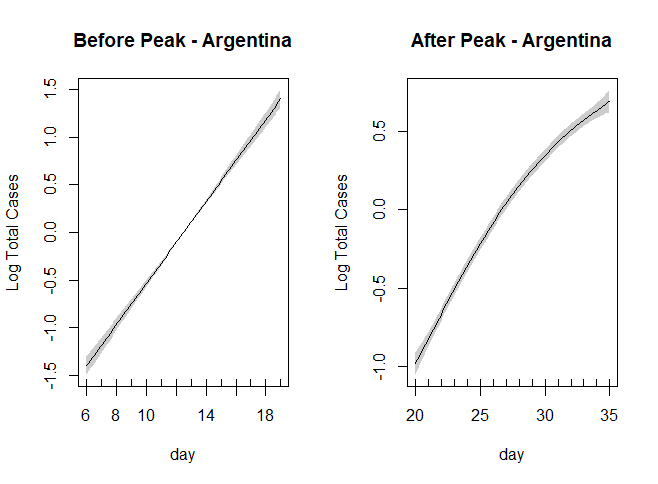
(B)
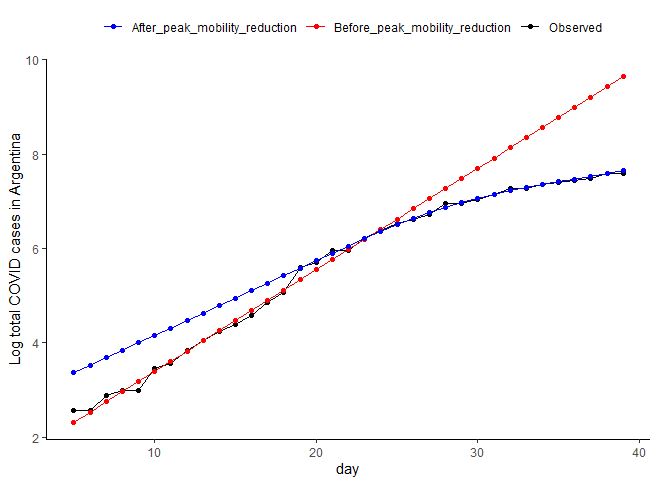

**Brazil**

(A)

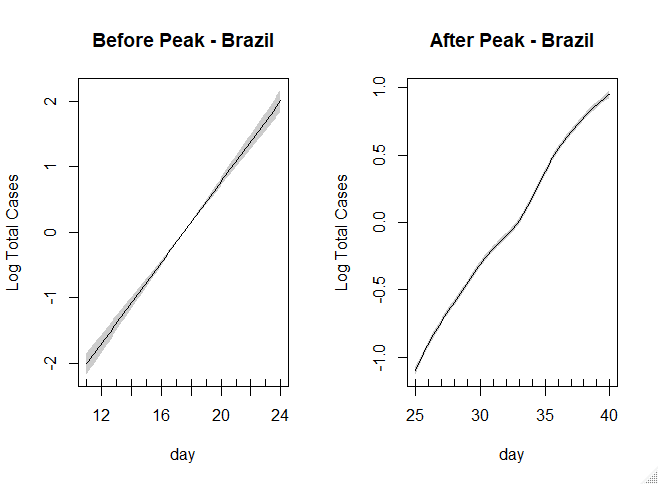
(B)
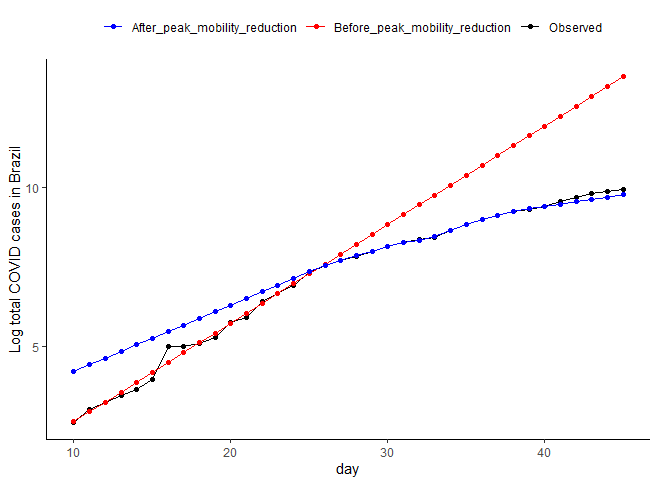

**Mexico**

(A)

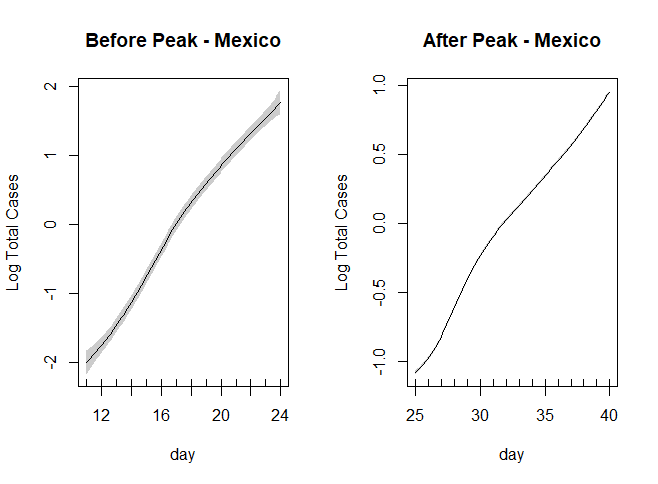
(B)
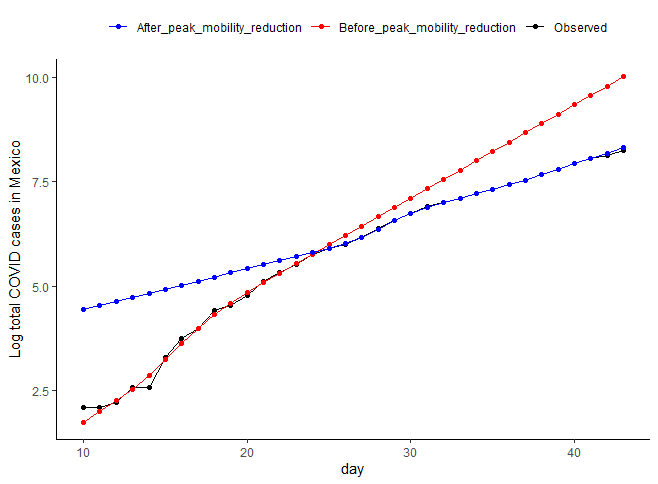

**Turkey**

(A)

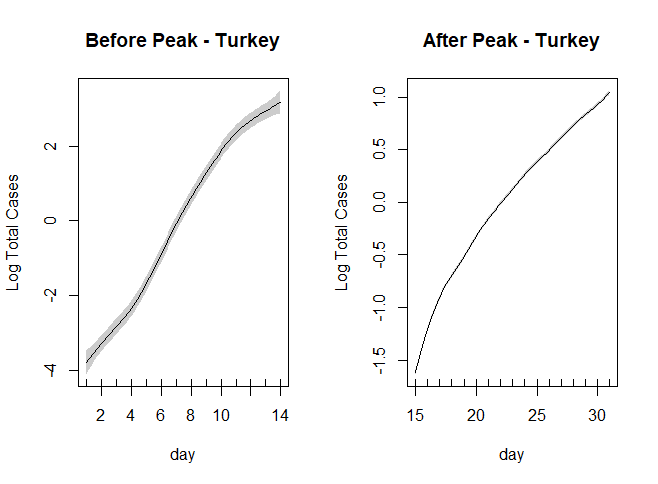
(B)

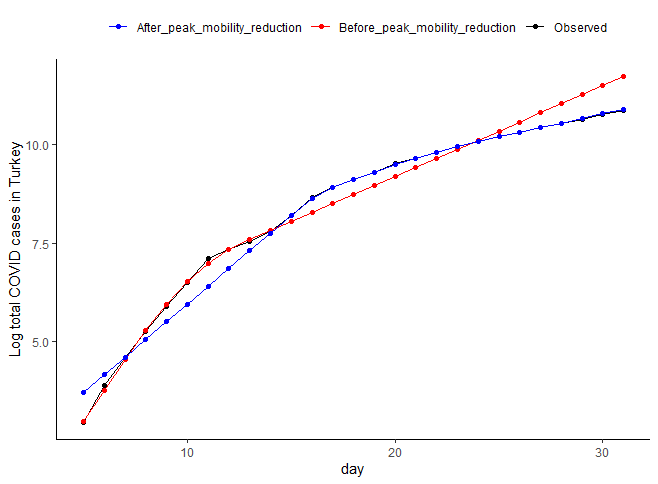

**Japan**

(A)

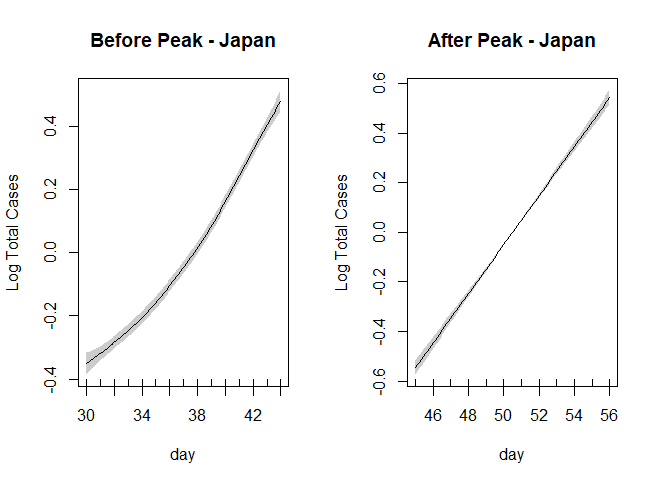

(B)

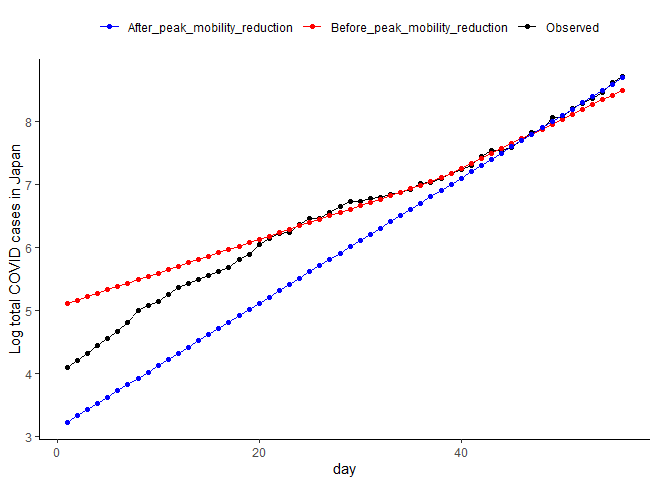

**Australia**

(A)

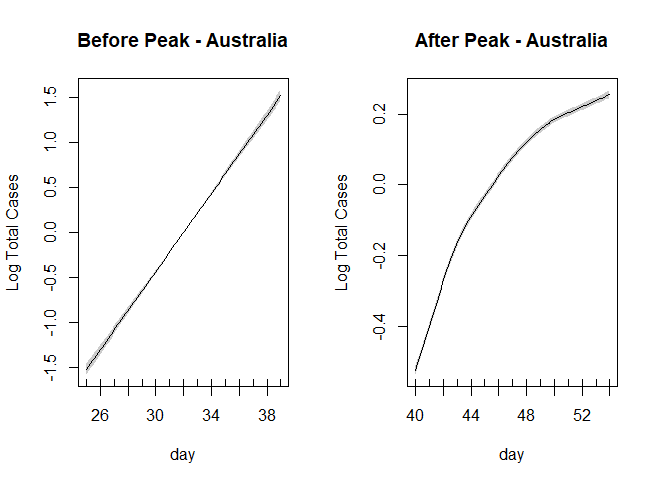

(B)

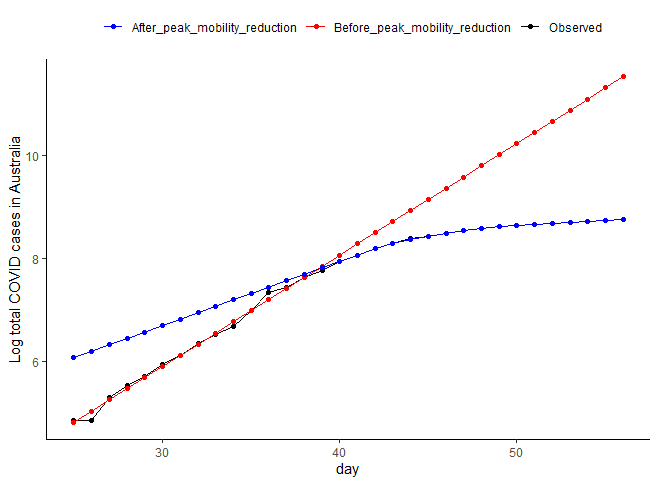

**Canada**

(A)

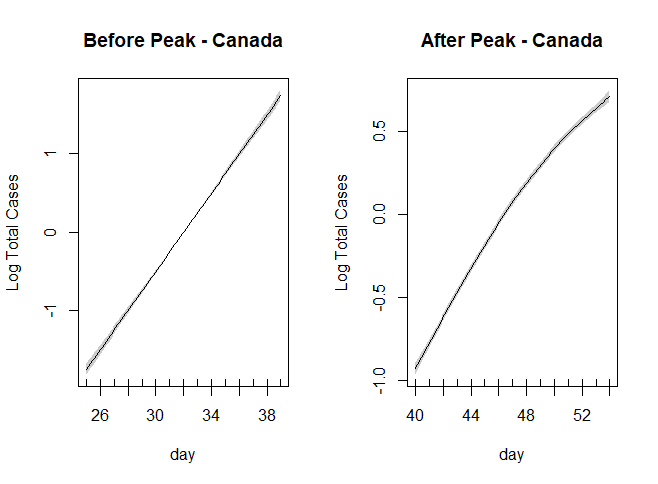

(B)

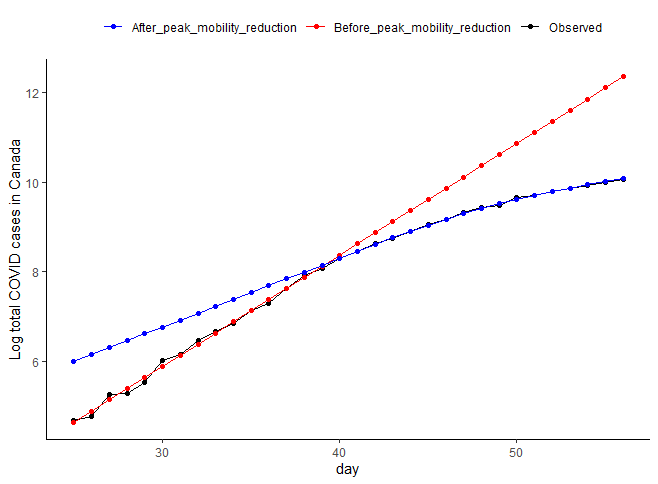

**South Korea**

(A)

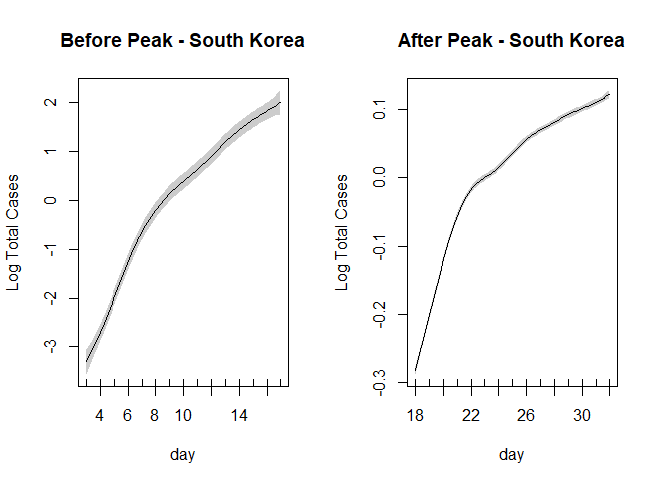

(B)

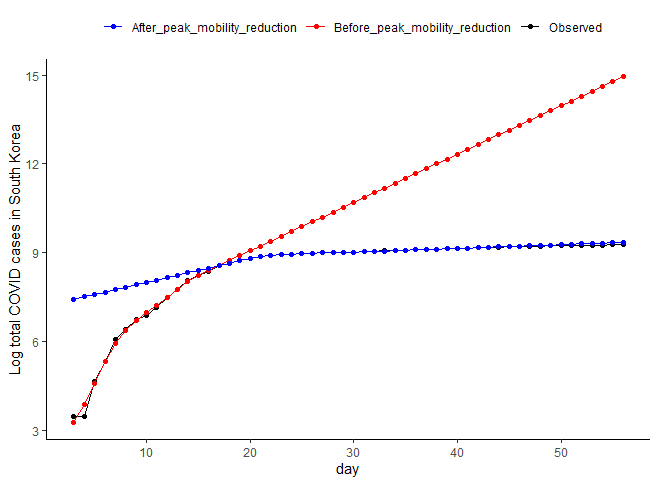

**Sweden**

(A)

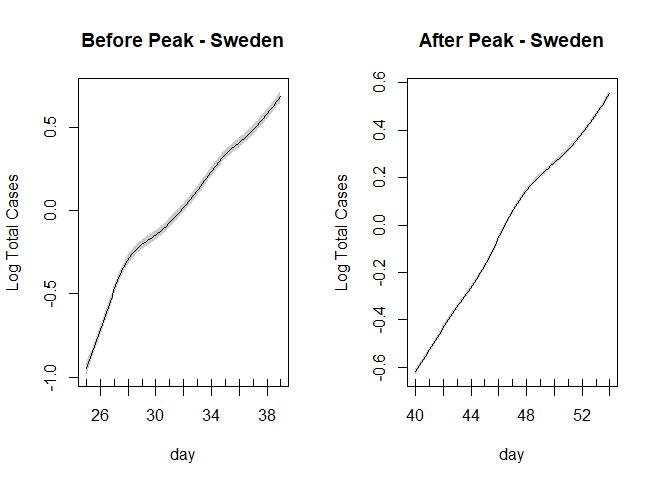
(B)

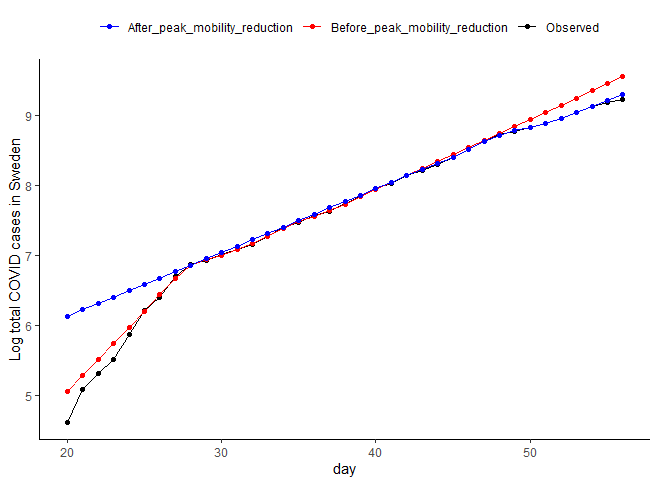

**United States**

(A)

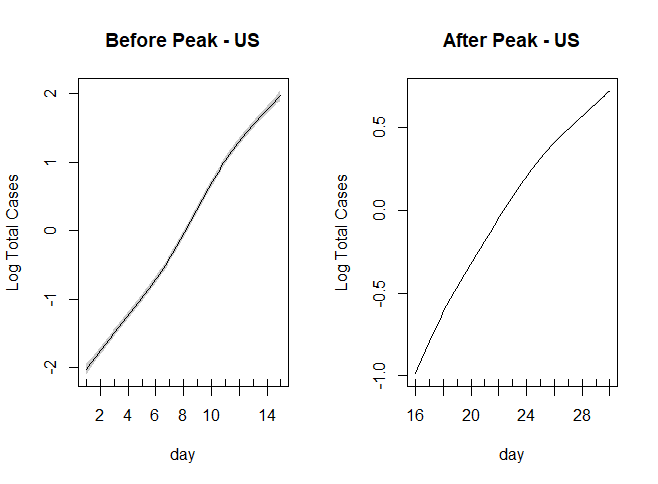

(B)

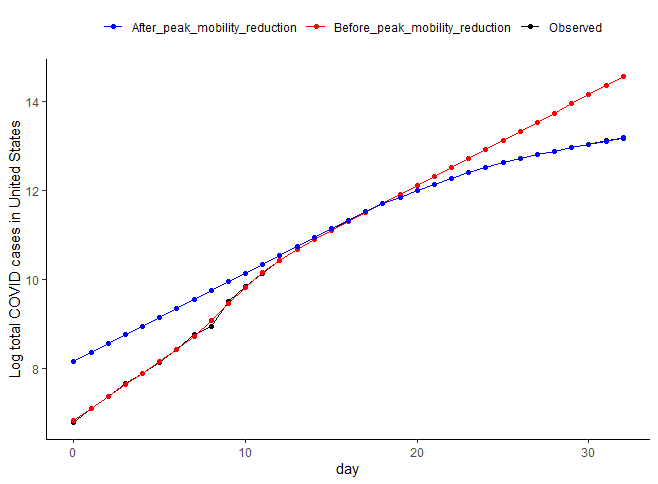
